# Longitudinal alterations in brain microstructure surrounding subcortical ischemic stroke lesions detected by free-water imaging

**DOI:** 10.1101/2023.04.14.23288593

**Authors:** Felix L. Nägele, Marvin Petersen, Carola Mayer, Marlene Bönstrup, Robert Schulz, Christian Gerloff, Götz Thomalla, Cheng Bastian

## Abstract

**Background:** Free-water imaging identifies subtle changes in white matter microstructure indicative of cellular and extracellular pathologies not visible on conventional stroke MRI. We explore the spatial extent and temporal trajectory of free-water changes in patients with subcortical stroke and their relationship to symptoms, as well as lesion evolution.

**Methods:** Twenty-seven patients with isolated subcortical infarct with mean age of 66.73 (SD 11.57) and median initial NIHSS score of 4 (IQR 4) received MRI 3-5 days, 1 month, 3 months and 12 months after symptom-onset. After lesion segmentation, 8 unique tissue shells (2 mm distance) surrounding stroke lesions were created. Extracellular freewater and fractional anisotropy of the tissue (FA_T_), derived from diffusion-weighted MRI, were averaged within tissue shells/stroke lesions, and normalized to corresponding contralateral regions. Linear mixed-effects models and t-tests were used for statistics. Baseline imaging measures were correlated with clinical outcomes 3 months after stroke.

**Results:** We found increased free-water and decreased FA_T_ in the stroke lesion, as well as the surrounding tissue with a characteristic spatio-temporal distribution. Free-water and FA_T_ changes were most prominent within the lesion and gradually became less with increasing distance from the lesion. Free-water elevations continuously increased over time and peaked after 12 months. In contrast, FA_T_ decreases were most pronounced 1 month after stroke, after which there was a steady increase leading to similarly reduced FA_T_ levels 12 months compared to 3-5 days after stroke. Higher perilesional free-water and higher lesional FA_T_ at baseline were correlated with greater reductions in lesion size, while there were no associations with clinical measures.

**Conclusions:** Both free-water and FA_T_ are altered beyond isolated subcortical stroke lesions. The spatial extent of these extracellular and cellular changes varies differentially over time indicating a dynamic parenchymal response to the initial insult characterized by vasogenic edema, cellular damage and white matter atrophy.

## INTRODUCTION

Ischemic stroke is among the leading causes of disability worldwide.^1^ Although lesion volume is a significant predictor of long-term outcomes^2^, it is well known that small, yet strategically located subcortical infarcts may just as well lead to debilitating impairments such as paresis^3,4^ or cognitive decline^5^.

While approximately 20-30 % of ischemic strokes are attributable to subcortical infarcts, their pathophysiology remains incompletely understood, not least because of the complex interplay between endothelial dysfunction, blood-brain barrier disruption, neuroinflammation and subsequent ischemic injury.^6,7^ In an effort to elucidate mechanisms of damage and repair which may ultimately be translated into specific treatment and prevention approaches, investigations of local tissue properties have recently gained attention.^6–8^ Neuroimaging has proven a powerful tool in furthering our understanding of *in-vivo* brain structural changes associated with ischemic stroke.^9,10^ In particular, diffusion-weighted magnetic resonance imaging (MRI) has been used extensively to describe degenerative alterations of white matter tracts following an ischemic event.^11^ Various diffusion tensor imaging (DTI) studies show that white matter degeneration occurs not only at the site of injury, but also in distant areas during the weeks after stroke.^12–16^ The latter phenomenon has been interpreted as imaging evidence of Wallerian degeneration and, in the case of the corticospinal tract, e.g., has been shown to be correlated with motor function.^16,17^ Whereas these tract-specific investigations provide plausible and important neuroanatomical explanations for structural brain changes and clinical outcomes, the role of the local tissue environment within and surrounding the primary ischemic lesion remains poorly understood.^18–22^

Recently, more advanced diffusion MRI models, such as free-water imaging, have been proposed to overcome methodological limitations of biological interpretability of conventional DTI metrics like fractional anisotropy (FA).^23,24^ Free-water imaging models two compartments: the first compartment reflects the volume fraction of isotropic diffusion of extracellular free-water at 37°C which may be sensitive to vasogenic edema or enlarged extracellular spaces due to atrophy.^23,25,26^ The second compartment models hindered/restricted diffusion in the proximity of cellular membranes of which FA of the tissue (FA_T_) can be estimated. Therefore, FAT more closely reflects the cellular microstructure of axons and myelin than conventional FA.^23^ While some studies^22,27–29^ have established free-water imaging as a useful tool to investigate stroke-related pathology, more fine grained longitudinal studies probing local cellular and extracellular changes in subcortical lesions and the surrounding tissue are lacking. However, such studies characterizing “penumbral” white matter alterations may provide important pathophysiological information which may ultimately be used to alter tissue fate and thus recovery trajectories in this relevant stroke population.

Utilizing free-water imaging, we therefore aimed to study cellular (FA_T_) and extracellular (free-water) changes within and surrounding subcortical stroke lesions from 3-5 days up to 12 months after the ischemic event in a well-defined longitudinal sample of subcortical stroke patients. Moreover, we evaluated associations of lesional and perilesional diffusion imaging markers with clinical measures, such as the National Institutes of Health Stroke Scale (NIHSS) and the Fugl-Meyer assessment of the upper extremity (UEFM), as well as with longitudinal change in stroke volume.

## METHODS

Reporting follows the STROBE (Strengthening the Reporting of Observational Studies in Epidemiology) guideline.^30^ The respective checklist is available in the **Supplementary Material**.

### Ethics

The current study was approved by the local ethics committee (Landesärztekammer Hamburg [State of Hamburg Chamber of Medical Practitioners]) and all participants gave written informed consent.

### Study population

We have previously reported on data of this longitudinal stroke cohort which was collected within the Collaborative Research Centre 936 between 2012 and 2017.^3,15^ Patients were recruited via the stroke unit of the University Medical Center Hamburg-Eppendorf 3–5 days after their first-time ischemic stroke in case they met the following inclusion criteria: (1) upper limb motor deficit; (2) isolated acute ischemic stroke lesion and absence of obvious white matter lesions or lesions from previous stroke determined by magnetic resonance imaging (MRI); (3) written informed consent; (4) absence of severe neurological or non-neurological co-morbidity. Exclusion criteria were conditions of pre-existing structural brain damage, e.g., previous stroke, intracranial hemorrhage, any other space-occupying lesion, as determined by fluid attenuated inversion recovery (FLAIR) and T1-weighted images. Upper limb motor deficits were defined as arm paresis or reduced finger dexterity as reported by the patient and confirmed in clinical examination. MRI and clinical examinations of patients were conducted at 4 time points: during the acute phase (3–5 days post-stroke), as well as in the subacute and chronic phases 30–40 days (∼ 1 month), 85–95 days (∼ 3 months), and 340–380 days (∼ 12 months) after stroke. For the current analysis, only patients with isolated subcortical infarcts were included.

### Clinical testing

Clinical examinations included the National Institute of Health Stroke Scale (NIHSS), the modified Rankin scale (mRS), the UEFM, nine hole peg test (NHP), as well as grip force of the affected hand (in kg), calculated as the mean of 3 consecutive measurements, relative to the unaffected hand.

### Image acquisition

Scanning was performed on a single 3T Siemens Skyra MRI scanner (Siemens, Erlangen, Germany) using a 32-channel head coil. Imaging sequences included highresolution T1-weighted anatomical, FLAIR and diffusion weighted images (DWI). For T1-weighted MRI, a 3D magnetization-prepared rapid acquisition gradient-echo sequence (MPRAGE) was employed with the following sequence parameters: repetition time (TR) = 2500 ms, echo time (TE) = 2.12 ms, field of view (FOV) = 240 x 192 mm, 256 axial slices, slice thickness (ST) = 0.94, in-plane resolution (IPR) = 0.94 x 0.94 mm. FLAIR images were acquired as follows: TR = 9000 ms, TE = 90 ms, TI = 2500 ms, FOV = 230 x 230 mm, ST = 5 mm, IPR = 0.7 x 0.7 mm. Finally, DWI included 75 axial slices with whole-brain coverage acquired along 64 non-collinear gradient directions (b = 1500 s/mm^2^) and one image with b = 0 s/mm^2^: TR = 10,000 ms, TE = 82 ms, FOV = 256 x 204, ST = 2 mm, IPR = 2 x 2 mm.

### Image processing

An overview of the imaging pipeline can be found in **Figure 1**. All code is publicly available on GitHub: https://github.com/csi-hamburg/CSIframe. After visual quality control of raw data, preprocessing of anatomical T1w and DWI was conducted in *QSIPrep* 0.14.2^31^ which is based on *Nipype* 1.6.1^32^. A more detailed description of image preprocessing can be found in the **Supplementary Material**.

**Figure 1.**
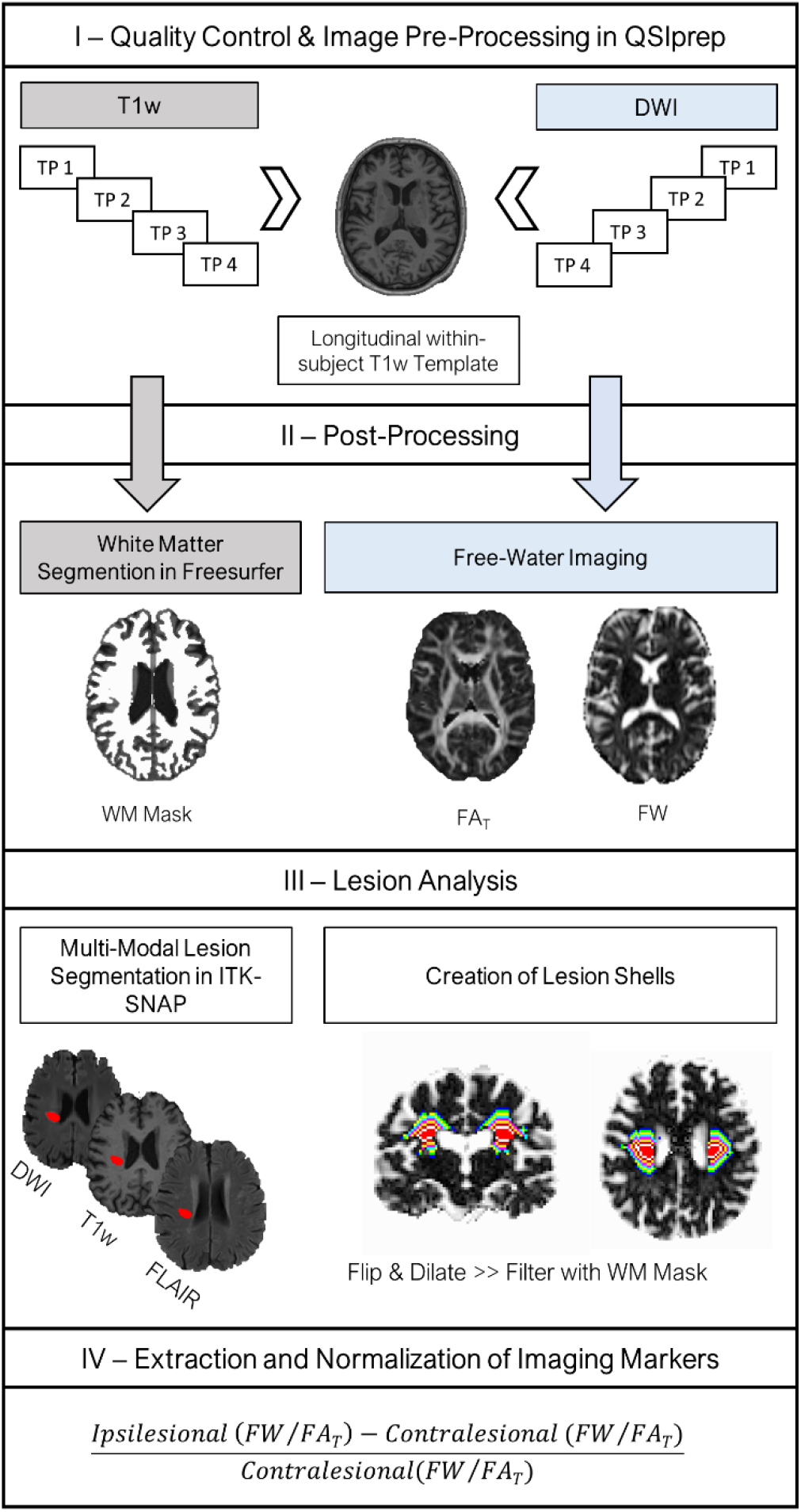
Overview of the imaging pipeline. Figure 1 schematically illustrates the basic steps of the imaging pipeline used for anaylsis. **(I)** First, standardized preprocessing of T1-weighted (T1w) and diffusion-weighted images (DWI), including but not limited to skull stripping, bias field and motion correction, was performed in QSIprep^31^. An unbiased longitudinal T1w within-subject template was created and subsequently used for co-registration of DWIs of each time point (3-5 days [TP1],1 month [TP2], 3 months [TP3] and 12 months after stroke [TP4]). **(II)** Next, a white matter mask based on the T1w-reference was generated with FreeSurfer for later processing steps. Moreover, the free-water bi-tensor model was fitted to the preprocessed DWI data to derive free-water and fractional anisotropy of the tissue (FA_T_) maps. **(III)** Stroke lesions were semi-automatically segmented in ITKSNAP based on FLAIR and T1w images, while DWI was used complementary to guide manual refinement of lesion masks. Lesion shells were created by iterative voxel-wise dilations of the lesion mask, resulting in 8 unique tissue shells. The same procedure was conducted after flipping the lesion to the contralateral side. Both ipsi- and contralateral tissue shells were filtered to include only white matter (WM) using the WM mask generated previously. **(IV)** Finally, average free-water and FA_T_ were extracted from the lesion and tissue shells and normalized to the corresponding contralateral region for further analysis.

#### Anatomical data preprocessing

We employed the longitudinal anatomical processing stream to ensure proper co-registration of individual time points for each participant. All T1w images were corrected for intensity non-uniformity (INU) using N4BiasFieldCorrection^33^ (ANTs 2.3.1). For each subject, a T1w-reference map was computed after registration of all T1w images/time points (after INU-correction) using mri_robust_template^34^ (FreeSurfer 6.0.1). The T1w-reference was then skull-stripped using antsBrainExtraction.sh (ANTs 2.3.1) and brain surfaces were reconstructed using recon-all^35^ (FreeSurfer 6.0.1).

#### Diffusion data preprocessing

First, denoising was performed with *MRtrix3*’s dwidenoise^36^. Next, Gibbs unringing was implemented using *MRtrix3*’s mrdegibbs^37^ after which, B1 field inhomogeneity was corrected using *MRtrix3*’s dwibiascorrect with the N4 algorithm^33^. *FSL*’s eddy was used for head motion correction and eddy current correction^38^ with outlier replacement^39^. Shells were aligned post-eddy. Further, susceptibility distortions were corrected based on *fMRIPrep*’s^40^ fieldmap-less approach^41,42^. Finally, the DWI time-series were resampled to ACPC, generating a preprocessed DWI run in ACPC T1w space with 2 mm isotropic voxels.

#### Free-water imaging

Free-water imaging, a two-tensor diffusion MRI model, was used to model an extracellular compartment of isotropic diffusion, as well as a cellular compartment characterized by hindered/restricted diffusion of water molecules.^23^ Thus, by means of a regularized non-linear fit of preprocessed DWIs, the volume fraction of free-water, and free-water corrected diffusion tensors were estimated at the voxel level for each study participant and time point. Finally, maps of fractional anisotropy of the tissue compartment (FA_T_) were derived based on the diffusion tensors.^23^

#### Lesion analysis

We performed multi-model semi-automatic lesion segmentation in *ITK-SNAP*^43^ (http://www.itksnap.org/) based on T1w and FLAIR signal alterations. DWI images were used complementary to guide the identification of stroke lesions. Lesions were segmented in T1w space with a clustering approach using linearly co-registered and preprocessed images of each modality. After the initial lesion masking, they were visually checked and manually edited by a research assistant to ensure accurate segmentations and coverage of visually apparent FLAIR and T1w signal alterations. Next, a second independent rater conducted visual quality control and editing. Deviations greater than 10% of the original lesion voxel size required a reconciliation with the person who prepared the initial lesion mask.

Further, for later normalization of free-water imaging markers, lesion masks were flipped to the contralateral hemisphere. This was conducted in standard space (MNI152NLin2009cAsym) to ensure anatomical correspondence. Flipped lesion masks were then back transformed to native T1w space for further processing. Both original and flipped lesion masks, as well as *FreeSurfer* white matter segmentations (derived through *QSIPrep*) were resampled to the DWI voxel size (2 x 2 x 2 mm). Next, 8 tissue shells surrounding the original and flipped lesion masks, separately, were created by iteratively dilating the masks (one iteration = 1 voxel, i.e., 2 mm) in three-dimensional space and subtracting the previous iterations from the dilated masks. This resulted in 2 x 8 unique tissue shells of increasing distance to the lesion masks with 2 mm width each. In an effort to reduce partial volume effects and to limit the analysis to the cerebral white matter, tissue shells were filtered with individual *FreeSurfer* white matter segmentations. Finally, we extracted average free-water and FA_T_ for each lesion and tissue shell and normalized the values to the corresponding contralateral, i.e. flipped, lesion/tissue shell for further statistical analysis (ipsilateral – contralateral / contralateral).

### Statistical analysis

All statistical analyses were conducted in R 4.1.3 (https://www.R-project.org/). Testing was performed two-tailed and the level of significance was set to p<.05. Missing data was not imputed.

#### Clinical data

Descriptive statistics of clinical characteristics at baseline and 3 months after stroke are presented as mean and standard deviation (SD) or absolute number and percentage.

#### Imaging

Our statistical analysis plan set out to answer 3 questions: (1) Do imaging markers in the lesion and tissue shells differ from the corresponding contralateral, healthy regions? (2) Are there differences in normalized imaging markers between the lesion and tissue shells? (3) Is there a longitudinal trend between time points in diffusion imaging markers across the lesion and tissue shells?

(1) To characterize the spatial extent of tissue alterations relative to healthy white matter, one-sample t-tests were performed with normalized diffusion markers for each region of interest and time point, separately. (2) For each time point, we used linear mixed effects models with *subject* as random factor to test for differences in normalized free-water and FA_T_ between regions of interest adjusting for *lesion volume* and *days from stroke to MRI session*. Helmert contrasts were selected to test for the main effect *location of measurement*, thus comparing each level with the mean of the subsequent levels. (3) Finally, we fitted linear mixed effects models with *subject* and *location of measurement* as random factors to test for differences in normalized freewater and FA_T_ across time points, again adjusting for lesion volume. Here, we used dummy coding for contrasts and performed Tukey post-hoc tests to compare time points with each other.

#### Correlation analysis

To interrogate associations between imaging measures and outcome variables, we performed Spearman correlations of baseline imaging markers in the lesion and tissue shells, separately, with NIHSS, relative grip force, UEFM and NHP at 3 months after stroke; as well as with relative change in lesion size from 3-5 days to 3 months after stroke.

## RESULTS

### Demographics and clinical characteristics

Descriptive statistics are presented in **Table 1**. DWI data from the original study was available for 52 patients. In total, 27 patients with subcortical ischemic stroke were included in the present analysis. Note that not all 27 participants completed all study visits (3-5 days: N = 26, 1 month: N = 21, 3 months: N = 19, 12 months: N = 19). The participants were on average 67 (SD 11.6) years old and presented with an average baseline NIHSS of 4.7 (SD 3.3). The sample was relatively well balanced regarding sex (41% female) and lesion side (44% left side). Numerically, all clinical outcome parameters improved from 3-5 days to 3 months after stroke, while lesion volumes decreased.

**Table 1.** Sample characteristics

| Variable* | Patients with subcortical ischemic stroke (N = 27) |  |
| --- | --- | --- |
| Age in years | 66.73 (11.57) |  |
| Female sex, N (%) | 11 (40.7 %) |  |
| Lesion side, left, N (%) | 12 (44.4 %) |  |
|  | 3-5 days after stroke | 3 months after stroke |
| Lesion volume in ml | 8.38 (11.89), 27 | 3.76 (5.53), 19 |
| NIHSS** | 4 (4), 25 | 1 (3), 19 |
| Relative grip strength of affected hand | 0.45 (0.40), 22 | 0.72 (0.40), 18 |
| UEFM | 37.92 (26.62), 24 | 50.79 (23.10), 19 |
| NHP | 2.10 (1.51), 12 | 1.24 (0.32), 14 |
| *If not specified otherwise, data are presented as mean (standard deviation [SD]), N<br>**Presented as median (inter-quartile range)<br>*Abbreviations:* NHP = Nine-Hole-Peg-Test, NIHSS = National Institutes of Health Stroke Scale, SD = standard deviation, UEFM = Fugl-Meyer assessment of the upper extremity |  |  |

### Imaging

#### Cross-sectional analyses comparing ipsilateral with contralateral tissue properties

Results of the cross-sectional analyses investigating differences between ipsilateral and contralateral tissue properties for each time point can be found in **Table 2**. Boxplots of imaging measures at the 4 different time points stratified by location of measurement are visualized in **Figure 2 (Panel A and B)**.

**Figure 2.**
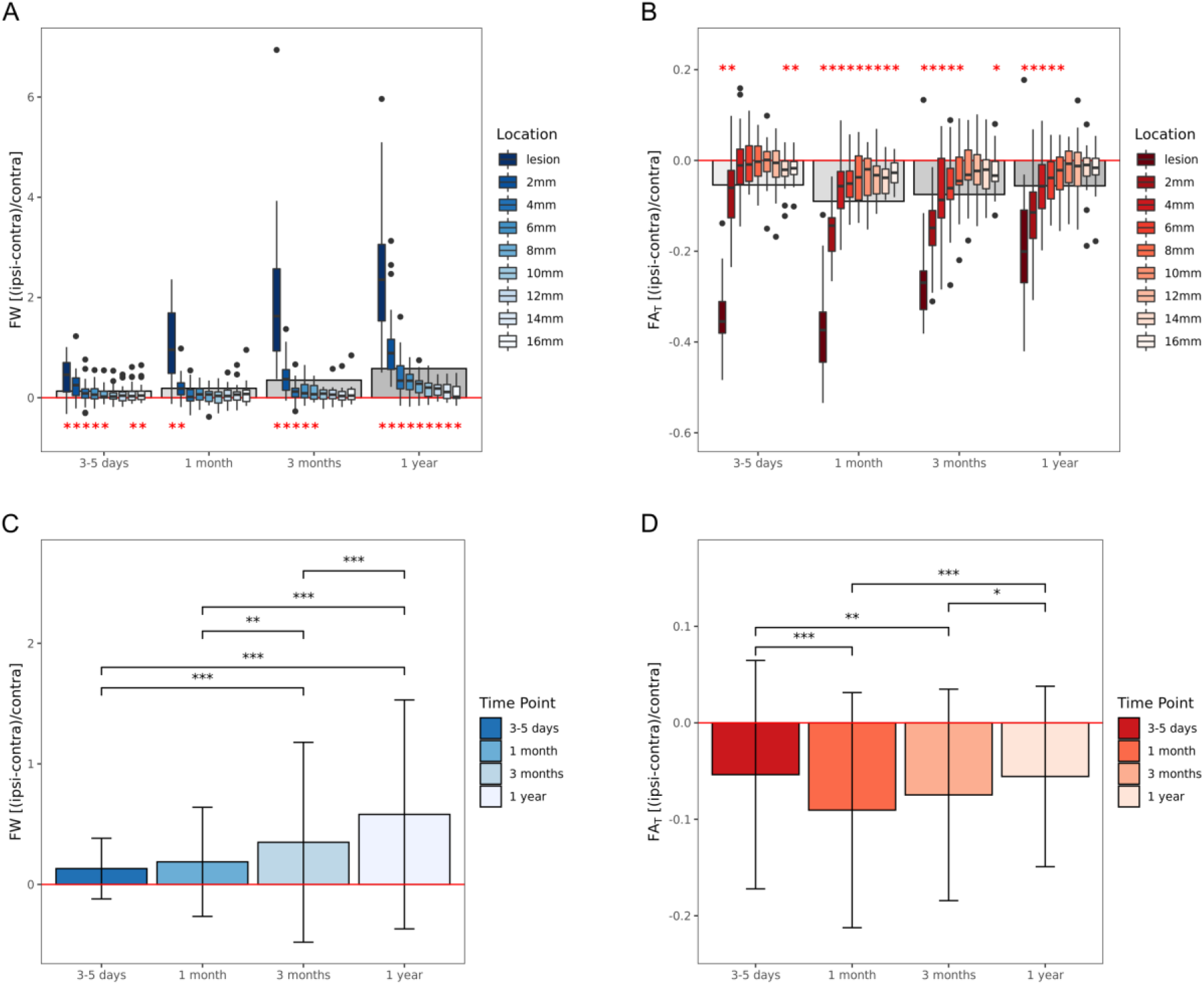
Box plots and bar graphs visualizing relative change in free-water and FA_T_ by time point and location of measurement. Panel A and B show the relative ispilateral change in free-water (A) and FA_T_ (B) compared to contralateral corresponding regions. The box plots are stratified by location of measurement and time point. The lower and upper end of the box corresonds to the 1^st^ and 3^rd^ quartiles, respectively. The line within the box represents the median. The whiskers extend to 1.5 times the interquartile range below and above the 1^st^ and 3^rd^ quartiles, respectively. The red horizontel line indicates the level of zero change, the gray rectangles in the background represent the mean across all regions of interest for each time point. Red asterisks indicate a significant difference from zero change (*P*<.05) as determined by one-sample t-tests for each region of interest, separately. **Panel C and D** show the relative ipsilateral change in free-water (C) and FA_T_ (D) averaged across regions of interest for each time point. The whiskers extend from one standard deviation above to one below the mean. Asterisks indicate the level of significance (\*\*\**P*<.001, \*\**P*<.01, \**P*<.05) derived from longitudinal linear mixed-effects models and post-hoc Tukey’s tests.

**Table 2.**
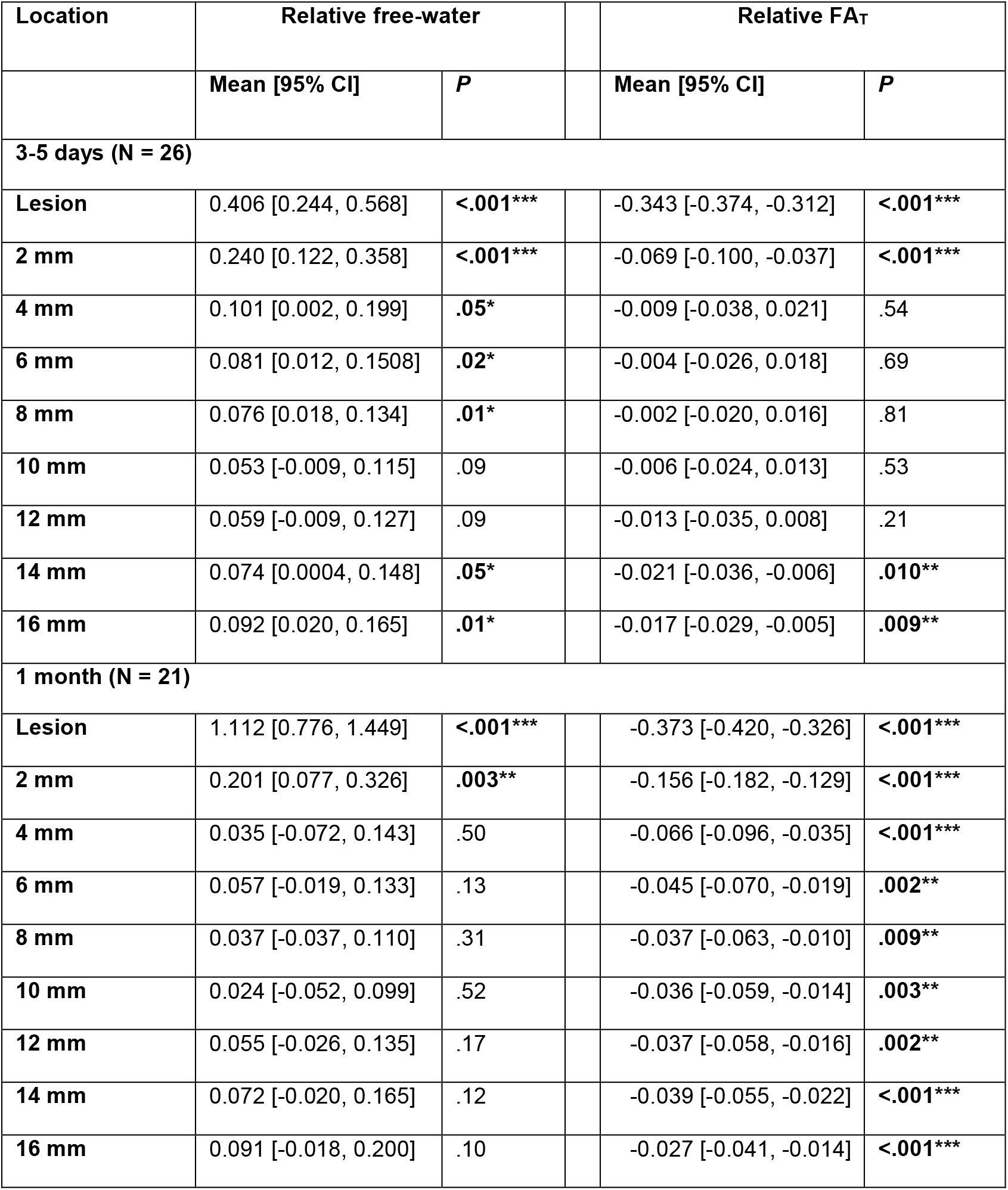
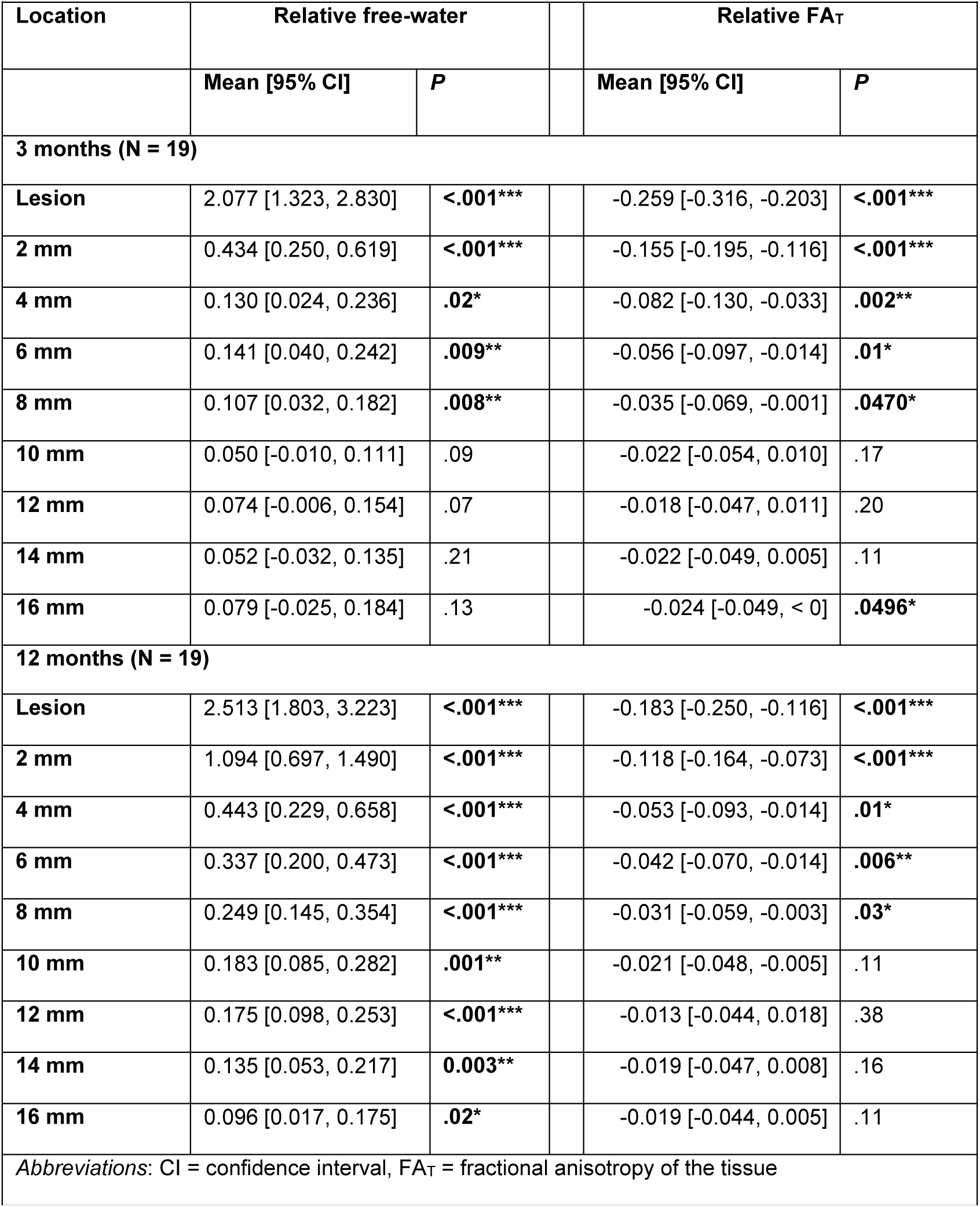
Results of one-sample t-tests for each time point, investigating whether ipsilesional diffusion parameters are different compared to corresponding contralateral regions.

As compared to healthy tissue, 3-5 days after stroke, free-water was significantly increased in the lesion, 2-8 mm, as well as 14-16 mm tissue shells. The most prominent increase compared to the contralesional hemisphere occurred in the lesion with +40.6% (*P*<.001), gradually decreasing to + 5.3% (*P*=.09) at 10 mm distance, increasing again to +9.2% (*P*=.01) at 16 mm distance. On the other hand, relative FA_T_ was decreased ipsilesionally comparing to contralesional healthy tissue. Significant reductions were found in the lesion (–34.3%, *P*<.001), at 2 mm (–6.9%, *P*<.001), 14 mm (–2.1%, *P*=.01) and 16 mm distance (–1.7%, *P*=.009).

One month after stroke, free-water alterations were less widespread with significant increases being only found in the lesion (+111.2%, *P*<.001) and 2 mm tissue shell (+20.1%, *P*=.003). In contrast, FA_T_, was significantly decreased in all regions of interest with a continuous gradient from –37.3% (P<.001) in the lesion to –2.7% (*P*<.001) at 16 mm distance.

At the third imaging assessment 3 months after stroke, significant free-water increases occurred in the lesion (+207.7%, *P*<.001) and at 2-8 mm distance ranging from +43.4% to +10.7% (*P*<.05), continuously decreasing with further distance. At the same time, FA_T_ decreases were less widespread comparing to 1 month after stroke. FA_T_ was significantly reduced by –25.9% in the lesion (*P*<.001), –15.5% at 2 mm (*P*<.001), –8.2% at 4 mm (*P*=.002), –5.6% at 6 mm (*P*=.01), –3.5% at 8 mm (*P*=.047) and by –2.4% at 16 mm distance (*P*=.0496).

Finally, at the last imaging examination 12 months after stroke, significant freewater alterations encompassed all regions of interest ranging from +251.3% increase in the lesion (*P*<.001) to +9.6% increase at 16 mm distance (*P*=.02). The spatial extent of FA_T_ reductions was comparable to the previous time point, i.e. FA_T_ was decreased by –18.3% in the lesion (*P*<.001), –11.8% at 2 mm (*P*<.001), –5.3% at 4 mm (*P*=.01), – 4.2% at 6mm (*P*=.006) and by –3.1% at 8 mm distance (*P*=.03).

#### Cross-sectional analyses comparing ipsilateral tissue properties between lesion and tissue shells

Detailed results of the linear mixed effects models investigating differences in relative free-water and FA_T_ between ipsilateral lesion and tissue shells can be found in the **Supplement** (**Table S1**).

Briefly, there was a significant main effect of *location of measurement* both in free-water and FA_T_ models at each time point (*P*<.001). Helmert contrasts revealed greater relative increases of free-water, as well as decreases of FA_T_ in the lesion and the 2 mm tissue shell compared to subsequent shells, respectively. One month after stroke, relative FA_T_ was additionally decreased at 4 mm distance comparing to shells with greater distance and there was a significant effect of *days since stroke* in the FA_T_ model (*β*=0.002, *P*=.02). Further at 3 months after stroke, baseline *lesion volume* predicted lower relative FA_T_ values (*β*=-0.006, *P*=.02).

#### Longitudinal analyses comparing ipsilateral relative free-water and FA_T_ between time points

Detailed results of the longitudinal linear mixed effects models testing for differences in relative free-water and FA_T_ between time points are shown in **Table 3** and visualized in **Figure 2 (Panel C and D)**.

**Table 3.** Results of the longitudinal linear mixed effects models investigating differences in free-water and FA_T_ between different time points

|  | Free-water |  |  | FA<sub>T</sub> |  |
| --- | --- | --- | --- | --- | --- |
|  | Estimate (SE) | P |  | Estimate (SE) | P |
| **Intercept** | 0.143 | .37 |  | -0.055 (0.031) | .12 |
| **Lesion volume<sup>a</sup>** | -0.001 (0.003) | .63 |  | <-0.001<br>(<0.001) | .60 |
| **Time point<sup>a</sup>** |  | **<.001\*\*\*** |  |  | **<.001\*\*\*** |
| **1 month – 3-5 days<sup>b</sup>** | 0.051 (0.050) | .74 |  | -0.035 (0.007) | **<.001\*\*** |
| **3 months – 3-5 days<sup>b</sup>** | 0.230 (0.052) | **<.001\*\*\*** |  | -0.025 (0.007) | **.003\*\*** |
| **1 year – 3-5 days<sup>b</sup>** | 0.455 (0.053) | **<.001\*\*\*** |  | -0.006 (0.007) | .82 |
| **3 months – 1 month<sup>b</sup>** | 0.179 (0.053) | **.004\*\*** |  | 0.010 (0.007) | .53 |
| **1 year – 1 month<sup>b</sup>** | 0.404 (0.053) | **<.001\*\*\*** |  | 0.029 (0.007) | **<.001\*\*\*** |
| **1 year – 3 months<sup>b</sup>** | 0.225 (0.053) | **<.001\*\*\*** |  | 0.019 (0.007) | **.04\*** |
| <sup>a</sup>Linear mixed-effects models with diffusion parameter as dependent variable, time point as fixed effect, and subject, as well as location as random intercept<br><sup>b</sup>Post-hoc Tukey's tests comparing different time points with one another<br>*Abbreviations:* FA<sub>T</sub> = fractional anisotropy of the tissue, SE = standard error |  |  |  |  |  |

*Time point* was a significant predictor of both relative free-water (*Χ^2^*=91.0(3), *P*<.001) and FA_T_ (*Χ^2^*=32.4(3), *P*<.001). Relative free-water increased continuously over time and was most prominent one year after stroke. Post-hoc Tukey tests revealed that all time points were significantly different from each other (*P*<.001), with the exception of 3-5 days compared to 1 month after stroke. The temporal dynamics of FA_T_ alterations were different compared to those of free-water. More specifically, relative FA_T_ decreases were most pronounced 1 month after stroke, followed by a steady increase, resulting in similarly reduced FA_T_ levels 12 months compared to 3-5 days after stroke.

#### Correlations

There were no significant associations between diffusion parameters (FW and FA_T_) 3-5 days after stroke and clinical outcomes 3 months later (**Table S2**). However, while 13 out of 18 (72%) lesions became smaller over time, we did observe a significant inverse correlation between perilesional free-water and change in lesion size, indicating that greater baseline free-water increases surrounding the lesion were associated with a greater reduction in lesion size after 3 months (*Rho*=-0.51, *P*=.03). In addition, higher lesional FA_T_ was significantly associated with greater reductions in lesion size after 3 months (*Rho*=-0.51, *P*=.03) (**Figure 3**).

**Figure 3.**
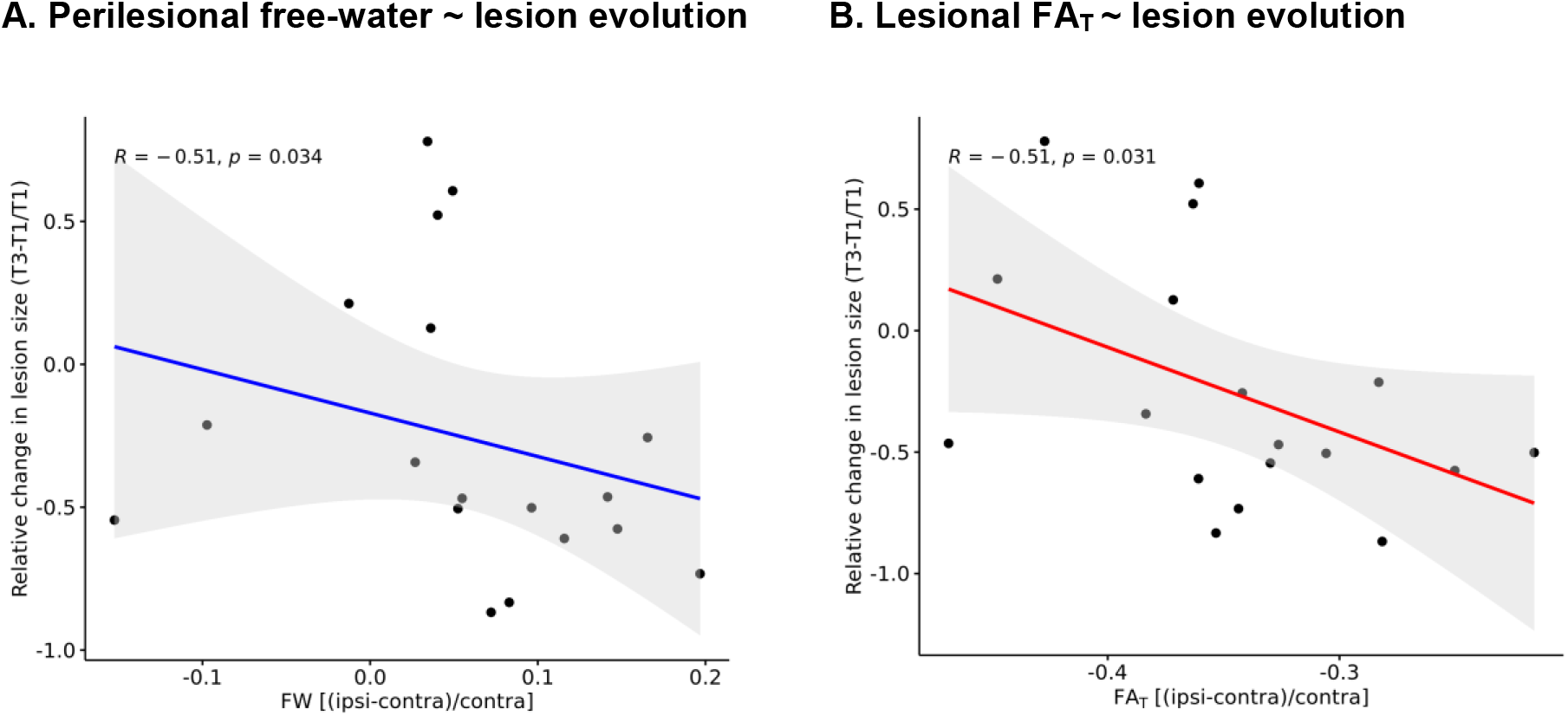
Correlations of perilesional free-water and lesional FA_T_ with lesion evolution. Panel A shows the significant negative correlation between mean relative *perilesional* free-water (FW) at baseline and relative change in lesion size between 3-5 days (T1) and 3 months (T3) after stroke. Increased perilesional free-water was significantly associated with greater reductions in lesion size within the first 3 months after stroke. **Panel B** illustrates the significant negative association of *lesional* fractional anisotropy of the tissue (FA_T_) and change in lesion size, indicating that higher FA_T_ is associated with greater reductions in lesion size.

## DISCUSSION

In a longitudinal free-water imaging study over 12 months after subcortical ischemic stroke, we identified dynamic changes of cellular and extracellular white matter alterations within the lesion and in brain tissue surrounding the primary stroke lesion. Compared to unaffected white matter on the contralesional hemisphere, free-water and FA_T_ were significantly altered in the lesion and the perilesional white matter at all investigated time points up to 16 mm distant to the original lesion. More specifically, freewater elevations – already evident at the initial assessment 3-5 days after stroke – continuously increased throughout the study period and were most prominent 12 months after stroke, affecting the entire surrounding tissue under investigation.

Cellular changes followed a different trajectory: FA_T_ decreases were most pronounced 1 month after stroke when they extended to all surrounding tissue shells, i.e., up to 16 mm distant to the lesion. Following this minimum, FA_T_ steadily increased again, reaching levels comparable to 3-5 days after stroke with remaining reductions in the lesion and up to 8 mm distance.

We did not observe significant correlations between baseline diffusion parameters and clinical outcomes 3 months after stroke. However, higher perilesional free-water and higher lesional FA_T_ were significantly associated with greater reduction in lesion size within the first 3 months after stroke.

### Free-water alterations

Our finding of increased extracellular free-water beyond visible lesions on conventional structural images, such as T1w and FLAIR images, is in line with previous reports on ischemic stroke patients in the acute to subacute^29^, as well as chronic phase^27,28^. Archer and co-workers^27^ assessed patients at least 6 months after stroke in a crosssectional, region of interest design and found increased relative free-water in the ipsi-lesional primary and premotor corticospinal tract. Similar to our study, Kern et al.^29^ employed a longitudinal approach comparing global free-water, averaged across the entire white matter, between 4 different time points (acute, 24 hours, 5 days and 1 month). Interestingly, they reported an initial free-water increase outside of the lesion within 24 hours, but did not show differences between 5 days and 1 month which is in line with the results of our longitudinal linear mixed effects model. However, our study extends this previous work in that we followed a more fine-grained approach and continued to map the trajectory up to 1 year after stroke. We found that free-water elevations relative to corresponding contralateral regions were present at each time point, although to a varying spatial extent with a progressive increase occurring subsequent to the 1-month assessment.

Various pathological mechanisms may be considered in explaining the observed lesional and perilesional extracellular free-water elevations. In the early stages of ischemic stroke, vasogenic edema develops in and around the infarct core, eventually becoming visible on T2-weighted FLAIR images within the first few hours after symptom onset.^44^ The free-water metric measures the amount of freely diffusing water molecules in the extracellular space and may thus be a sensitive marker of vasogenic edema due to upregulated immune responses and consecutive blood-brain-barrier leakage.^23,25,45,46^ It is therefore conceivable that the early increases in extracellular diffusivity, which we observe in and beyond the lesion 3-5 days after stroke, are attributable to vasogenic edema and inflammatory processes.^47,48,29^ These free-water increases seem to diminish in the surrounding tissue within the first month, suggesting the regredience of edema in this time period. On the other hand, consistent with previous imaging and preclinical histopathological studies^15,27,46,22^, we observed free-water increases 3 and 12 months after stroke that are most likely explained by secondary mechanisms of neurodegeneration at the microscopic level such as fluid-filled cavitation and axonal loss (i.e., atrophy) leading to larger extracellular spaces. Intriguingly, these changes were also observed in the normal appearing white matter surrounding the stroke lesion. While our findings fit well with previous work, larger studies which include immune biomarker sampling, as well as the application of diffusion models that account for a vascular compartment^49^ and/or perfusion MRI could strengthen our understanding of the observed spatial and temporal trajectory of free-water alterations.

### FA_T_ alterations

Compared to free-water, we observed a different temporal trajectory for FA_T_ suggestive of diverging pathophysiological processes. Similar to free-water, FA_T_ was altered extending to regions well beyond the visible lesion. The detected reductions in white matter FA_T_ are in line with previous free-water imaging studies assessing cellular microstructure of the corticospinal tract.^27,28^ In a previous study, no longitudinal changes in whole-brain FA_T_ were observed, leading the authors to conclude that FA/FA_T_ might rather capture underlying brain health than stroke-associated changes.^29^ In contrast, in our sample, relative FA_T_ further decreased from 3-5 days to 1 month after stroke, before returning close to baseline levels after 1 year. These changes may be refined to tissue alterations surrounding the lesion und thus escape a global approach using a whole brain analysis.

Our finding of decreased FA_T_ despite co-occurring increases in free-water suggests that reductions in diffusion anisotropy in the proximity of cellular membranes are likely related to damage of myelin sheaths and/or axonal membranes in the cerebral white matter, as previously reported.^50^ Further, these tissue alterations occurred not only in the lesion, but extended to regions of 16 mm distance, in line with the concept of Wallerian degeneration.^16,51,52^ Interestingly, FA_T_ reductions compared to the unaffected hemisphere were most prominent 1 month after stroke, followed by a relative “recovery” during the following months.

Different potential mechanisms may explain this somewhat surprising trajectory. Initial structural damage caused by ischemia and the following Wallerian degeneration leads to disintegration of axons and surrounding myelin^53^ indicated by reductions in FA_T_. Thereafter, clearance of cellular debris and the accompanying atrophy of white matter tracts may lead to larger extracellular spaces (as evidenced by free-water elevations), reduced populations of crossing fibers and a denser configuration of the remaining axons, potentially alleviating FA_T_ reductions.^15,53^. Corroborating this hypothesis, a recent report by our group showed a trend of increasing fiber density – a fixel-based imaging metric^24^ – in the ipsilateral corticospinal tract of an overlapping sample (including also non-subcortical stroke patients) between 1 month and 1 year poststroke.^15^ Lastly, the formation of dense glial scars^53–55^ observed several months after cerebral infarction may contribute to higher diffusion anisotropy^56^ compared to earlier time points. In summary, we observed *in-vivo* evidence of dynamic changes in the lesional and perilesional cellular tissue compartment which are in agreement with established concepts in stroke research. Nevertheless, more research employing similar and different imaging methodologies are needed to further our understanding of the suggested mechanisms.

### Correlations of imaging measures with clinical outcomes

In our sample of subcortical stroke patients with upper limb motor deficit, we did not observe significant associations of baseline free-water imaging metrics in the lesion or surrounding tissue with clinical outcomes after 3 months. We believe that the discrep-ancy to earlier studies^27–29^ might be due to methodological differences. While our distance-dependent region of interest approach provided specificity in terms of spatial relation to the lesion, white matter regions not primarily involved in motor networks were included in our analysis, plausibly obscuring structure ∼ (motor) function relationships previously shown in tract-specific study designs. Moreover, the sample size, especially in the longitudinal analysis, was modest which likely contributed to a lack of power.

On the other hand, similar to a recent study by Duering and colleagues^22^, which reported on within-lesion heterogeneity of free-water imaging metrics in terms of subsequent tissue fate, we did observe preliminary evidence that free-water and FA_T_ were associated with the evolution of lesion size. Yet, in contrast to our study, Duering et al.^22^ did not investigate perilesional changes, but found a trend of lower lesional freewater in areas that appeared normal on conventional MRI at follow-up. Considering the explorative nature of our correlation analyses, as well as the modest sample size, more research is necessary to establish the clinical significance of altered lesional and perilesional tissue properties detected by diffusion MRI.

### Strengths and limitations

Strengths of our study include (1) a well-defined subcortical stroke population, (2) a state-of-the-art, well-documented and reproducible image processing pipeline, (3) an innovative, fine grained approach to study *in-vivo* white matter tissue properties surrounding stroke lesions, and (4) the longitudinal study design, allowing us to study the longer term dynamics of perilesional microstructural changes. Nevertheless, several limitations need to be mentioned. While focusing on subcortical stroke may have improved the sensitivity to detect common changes among patients, the generalizability to larger, territorial stroke subtypes remains to be established. Further, as mentioned above, we acknowledge that the sample size was small, and emphasize the need for larger replication studies. Lastly, our approach does not provide tract-specific information, and may therefore be limited its ability to elucidate brain-behavior relationships.

### Summary

Utilizing free-water imaging and a perilesional region of interest approach, our longitudinal study provides novel evidence of dynamic differential extracellular and cellular tissue alterations within *and* beyond subcortical stroke lesions visible on conventional MRI. Mapping the trajectories of these changes, we find that extracellular free-water elevations, potentially indicative of neuroinflammation and vasogenic edema in the acute phase, decline within the first month, before continuously increasing again in the chronic phase, suggestive of secondary white matter damage. In contrast, FA_T_ reductions peaked in the sub-acute phase (i.e., after 1 month), indicating that most axonal and myelin damage has occurred by then, whereas later stages may be characterized by microstructural reorganisation. External validation of our findings in larger samples including different types of ischemic stroke are needed.

## Data Availability

All data produced in the present study are available upon reasonable request to the authors

## ACKNOWLEDGMENTS

We describe contributions to the paper using the CRediT contributor role taxonomy. Conceptualization: F.L.N., G.T., B.C. Data Curation: F.L.N, M.P., C.M.; Formal analysis: F.L.N.; Investigation: F.L.N., M.P., C.M., M.B., R.S., C.G., G.T., B.C.; Methodology: F.L.N., C.M.; Software: F.L.N., M.P., C.M.; Supervision: G.T., B.C.; Funding Acquisition: C.G., G.T., B.C.; Visualization: F.L.N.; Writing—original draft: F.L.N.; Writing—review & editing: F.L.N., M.P., C.M., M.B., R.S., C.G., G.T., B.C. The authors declare no competing interests.

