## Supplementary Material for "Longitudinal alterations in brain microstructure surrounding subcortical ischemic stroke lesions detected by free-water imaging"

#### Methods

##### Image Processing

The following description is based on a boilerplate generated by *QSIPrep* and therefore facilitates standardized reporting and reproducibility.

*Anatomical data preprocessing*

We employed the longitudinal anatomical processing stream in order to ensure proper co-registration of individual time points for each participant. All T1w images were corrected for intensity non-uniformity (INU) using N4BiasFieldCorrection[1] (ANTs 2.3.1). For each subject, a T1w-reference map was computed after registration of all T1w images/time points (after INU-correction) using mri_robust_template[2] (FreeSurfer 6.0.1). The T1w-reference was then skull-stripped using antsBrainExtraction.sh (ANTs 2.3.1), using OASIS as target template. Brain surfaces were reconstructed using recon-all[3] (FreeSurfer 6.0.1), and the brain mask estimated previously was refined with a custom variation of the method to reconcile ANTs-derived and FreeSurfer-derived segmentations of the cortical gray-matter of Mindboggle[4].

*Diffusion data preprocessing*

MP-PCA denoising as implemented in MRtrix3's dwidenoise[5] was applied with a 5-voxel window. After MP-PCA, Gibbs unringing was performed using MRtrix3's mrdegibbs[6]. Following unringing, B1 field inhomogeneity was corrected using dwibiascorrect from MRtrix3 with the N4 algorithm[1].

*FSL*'s eddy (version 6.0.3:b862cdd5) was used for head motion correction and eddy current correction[7]. Eddy was configured with a *q*-space smoothing factor of 10, a total of 5 iterations, and 1000 voxels used to estimate hyperparameters. A linear first level model and a linear second level model were used to characterize eddy current-related spatial distortion. *q*-space coordinates were forcefully assigned to shells. Field offset was attempted to be separated from subject movement. Shells were aligned post-eddy. Eddy's outlier replacement was run[8]. Data were grouped by slice, only including values from slices determined to contain at least 250 intracerebral voxels. Groups deviating by more than 4 standard deviations from the prediction had their data replaced with imputed values. Final interpolation was performed using the jac method.

A deformation field to correct for susceptibility distortions was estimated based on *fMRIPrep*'s[9] fieldmap-less approach. The deformation field is that resulting from co-registering the b0 reference to the same-subject T1w-reference with its intensity inverted.[10,11] Registration is performed with antsRegistration (ANTs 2.3.1), and the process regularized by constraining deformation to be nonzero only along the phase-encoding direction, and modulated with an average fieldmap template[12]. Based on the estimated susceptibility distortion, an unwarped b=0 reference was calculated for a more accurate co-registration with the anatomical reference. The DWI time-series were resampled to ACPC, generating a preprocessed DWI run in ACPC T1w space with 2 mm isotropic voxels.

Many internal operations of *QSIPrep* use *Nilearn* 0.8.0[13] and *Dipy*[14]. For more details of the pipeline, see https://qsiprep.readthedocs.io/en/latest/workflows.html).

### Results

**Table S1. Results of the linear mixed effects models investigating differences in free-water and $FA_T$ between lesion and tissue shells at four different time points**

| | Free-water | | $FA_T$ | |
|---|---|---|---|---|
| | Estimate (SE) | *P* | Estimate (SE) | *P* |
| **Days 3-5 (N = 26)** | | | | |
| **Intercept** | 0.006 (0.129) | .96 | -0.043 (0.030) | .17 |
| **Location** | | | | |
| **Lesion** | 0.309 (0.041) | **<.001*** ** | -0.325 (0.011) | **<.001*** ** |
| **2 mm** | 0.163 (0.041) | **<.001*** ** | -0.058 (0.011) | **<.001*** ** |
| **4 mm** | 0.028 (0.041) | .50 | 0.002 (0.011) | .88 |
| **6 mm** | 0.010 (0.042) | .82 | 0.007 (0.011) | .49 |
| **8 mm** | 0.007 (0.043) | .88 | 0.012 (0.011) | .28 |
| **10 mm** | -0.022 (0.044) | .62 | 0.011 (0.011) | .33 |
| **12 mm** | -0.024 (0.047) | .60 | 0.005 (0.012) | .66 |
| **14 mm** | -0.018 (0.054) | .74 | -0.004 (0.014) | .78 |
| **Lesion volume** | 0.001 (0.002) | .55 | <-0.001 (<0.001) | .73 |
| **Days since stroke** | 0.029 (0.032) | .38 | -0.002 (0.007) | .76 |
| **1 month (N = 21)** | | | | |
| **Intercept** | 0.263 (0.157) | .11 | -0.151 (0.030) | **<.001*** ** |
| **Location** | | | | |
| **Lesion** | 1.041 (0.069) | **<.001*** ** | -0.318 (0.012) | **<.001*** ** |
| **2 mm** | 0.148 (0.070) | **.04*** | -0.115 (0.012) | **<.001*** ** |
| **4 mm** | -.021 (0.071) | .77 | -0.029 (0.013) | **.02*** |
| **6 mm** | 0.001 (0.072) | .99 | -0.010 (0.013) | .45 |
| **8 mm** | -0.024 (0.073) | .75 | -0.002 (0.013) | .88 |
| **10 mm** | -0.049 (0.076) | .52 | -0.002 (0.013) | .87 |
| **12 mm** | -0.027 (0.080) | .73 | -0.004 (0.014) | .79 |
| **14 mm** | -0.019 (0.093) | .84 | -0.011 (0.016) | .49 |
| **Lesion volume** | 0.002 (0.004) | .59 | -0.001 (0.001) | .20 |
| **Days since stroke** | -0.002 (0.003) | .54 | 0.002 (0.001) | **.02*** |

**Table S1. (continued)**

| | Free-water | | FA$_T$ | |
|---|---|---|---|---|
| | Estimate (SE) | *P* | Estimate (SE) | *P* |
| **3 months (N = 19)** | | | | |
| **Intercept** | 0.079 (0.311) | .80 | -0.072 (0.056) | .22 |
| **Location** | | | | |
| **Lesion** | 1.943 (0.132) | **<.001*** ** | -0.208 (0.016) | **<.001*** ** |
| **2 mm** | 0.344 (0.133) | **.01*** | -0.119 (0.016) | **<.001*** ** |
| **4 mm** | 0.046 (0.135) | .73 | -0.052 (0.016) | **.001** ** |
| **6 mm** | 0.069 (0.137) | .62 | -0.031 (0.016) | 0.05 |
| **8 mm** | 0.043 (0.139) | .76 | -0.013 (0.017) | .42 |
| **10 mm** | -0.018 (0.144) | .90 | <-0.001 (0.017) | .99 |
| **12 mm** | 0.008 (0.153) | .96 | 0.005 (0.018) | .79 |
| **14 mm** | -0.028 (0.176) | .88 | 0.003 (0.021) | .90 |
| **Lesion volume** | -0.003 (0.010) | .76 | -0.006 (0.002) | **.005** ** |
| **Days since stroke** | 0.003 (0.003) | .37 | <0.001 (0.001) | .74 |
| **12 months (N = 19)** | | | | |
| **Intercept** | 2.343 (1.720) | .19 | -0.024 (0.195) | .90 |
| **Location** | | | | |
| **Lesion** | 2.174 (0.133) | **<.001*** ** | -0.143 (0.018) | **<.001*** ** |
| **2 mm** | 0.862 (0.134) | **<.001*** ** | -0.090 (0.018) | **<.001*** ** |
| **4 mm** | 0.247 (0.136) | .7 | -0.030 (0.018) | .11 |
| **6 mm** | 0.169 (0.137) | .22 | -0.021 (0.018) | .25 |
| **8 mm** | 0.102 (0.140) | .47 | -0.012 (0.019) | .51 |
| **10 mm** | 0.048 (0.145) | .74 | -0.004 (0.019) | .83 |
| **12 mm** | 0.060 (0.154) | .70 | 0.006 (0.019) | .75 |
| **14 mm** | 0.039 (0.177) | .83 | <0.001 (0.023) | >.99 |
| **Lesion volume** | 0.011 (0.013) | .41 | -0.003 (0.001) | .06 |
| **Days since stroke** | -0.005 (0.005) | .31 | <0.001 (0.001) | .91 |

**Table S2. Spearman correlations of imaging parameters 3-5 days after stroke with clinical variables and change in lesion size 3 months after stroke**

| | Free-water | | | | $FA_T$ | | | |
|---|---|---|---|---|---|---|---|---|
| **Outcome variable** | **Lesional** | | **Perilesional** | | **Lesional** | | **Perilesional** | |
| | ***Rho*** | ***P*** | ***Rho*** | ***P*** | ***Rho*** | ***P*** | ***Rho*** | ***P*** |
| **Change in lesion size (N = 18)** | 0.14 | .59 | **-0.51** | **.03*** | **-0.51** | **.03*** | -0.09 | .71 |
| **NIHSS (N = 18)** | -0.29 | .24 | 0.18 | .47 | -0.15 | .55 | -0.08 | .74 |
| **Relative grip strength (N = 17)** | 0.21 | .42 | -0.22 | .40 | -0.04 | .89 | -0.06 | .82 |
| **UEFM (N = 18)** | 0.18 | .47 | 0.05 | .85 | 0.19 | .46 | -0.15 | .56 |
| **NHP (N = 13)** | 0.45 | .12 | 0.49 | .09 | 0.02 | .96 | -0.22 | .47 |

*Abbreviations*: $FA_T$ = fractional anisotropy of the tissue, NHP = Nine-Hole-Peg-Test, NIHSS = National Institutes of Health Stroke Scale, SD = standard deviation, UEFM = Fugl-Meyer assessment of the upper extremity
